# Novel blood protein biomarkers for distinguishing bacterial from non-bacterial infection in children: a case–control study

**DOI:** 10.1101/2025.10.17.25338235

**Authors:** Holly Drummond, Cathal Roarty, Helen Groves, Thomas Waterfield, Clare Mills

## Abstract

**Background:** Distinguishing bacterial from viral or inflammatory conditions in hospitalised children is clinically challenging due to overlapping clinical features and limitations of existing diagnostics. Culture-based tests are slow, and commonly used biomarkers such as C-reactive protein (CRP) lack sufficient accuracy, contributing to empirical antibiotic use and antimicrobial resistance. We aimed to evaluate the diagnostic performance of novel blood protein biomarkers for bacterial infection in children admitted with infection or inflammatory illness.

**Methods:** We performed a prospective case–control study of children ≤16 years admitted to the Royal Belfast Hospital for Sick Children (2020–2023) with bacterial infection, viral infection or inflammatory illness. Plasma samples were analysed for previously reported novel biomarkers and signatures (LCN2; TRAIL+IP-10+CRP; E-selectin+IL18+ NCAM1+LCN2+IFN-γ+LG3BP and E-selectin+IL18+ NCAM1+LCN2+IFN-γ). The diagnostic accuracy of individual biomarkers and biomarker signatures was assessed using ROC curves.

**Results:** Fifty-two children were included (13 bacterial, 20 viral, 19 inflammatory). All evaluated biomarker signatures and LCN2 distinguished bacterial from viral infection (AUC 0.819– 0.935), with the TRAIL+IP-10+CRP signature achieving the highest accuracy (AUC 0.935). In bacterial–inflammatory comparisons, performance was lower (AUCs 0.607–0.745); the E-selectin+IL-18+NCAM1+LCN2+IFN-γ signature performed best (AUC 0.745) and outperformed CRP (AUC 0.595). A novel three-protein signature (E-selectin+LCN2+IFN-γ) had a significantly higher AUC than CRP for distinguishing bacterial infections from non-bacterial (viral and inflammatory combined).

**Conclusions:** Several novel host protein biomarkers and signatures had a higher diagnostic accuracy than CRP for differentiating bacterial from non-bacterial illness (viral and inflammatory) in hospitalised children. These findings support the potential of biomarker-guided diagnostics to improve accuracy and facilitate earlier antibiotic de-escalation.

## Introduction

Infections in hospitalised children, which can be community-acquired or nosocomial, are a common cause of mortality and morbidity. Frequent community-acquired bacterial infections that often result in hospitalisation include pneumonia, urinary tract infections (UTIs) and bacteraemia (1). Nosocomial infections, often associated with the use of invasive devices and procedures, similarly include bacteraemia, pneumonia, and UTIs (2, 3).

Despite the high prevalence of infections in hospitalised children, accurately identifying infection aetiology remains challenging due to overlapping clinical features between bacterial, viral and non-infectious inflammatory conditions. Bacterial culture-based tests from sterile sites are slow, often taking days, limiting their use for initial antibiotic decision-making (4). Rapid molecular tests are limited by high pathogen carriage in children and restricted panel coverage (5). Although host biomarkers C-reactive protein (CRP) and procalcitonin (PCT) are widely used and validated in distinguishing bacterial from non-bacterial infections, multiple studies report their suboptimal accuracy (6, 7).

Given the limitations of current diagnostics, clinicians often take a cautious approach to antibiotic use in hospitalised children, whose immune defences are frequently impaired due to acute illness or underlying comorbidities (2). In children hospitalised in the paediatric intensive care unit, data suggests that antimicrobial utilisation can reach approximately 70% (8). However, inappropriate antibiotic use drives antimicrobial resistance, a growing public health threat (9). Given the severity of illness in this population, withholding antibiotics initially is rarely feasible; however, using biomarkers to guide antibiotic de-escalation offers a more practical strategy (10). Several promising biomarkers and biomarker signatures with improved diagnostic accuracy over CRP and PCT have been reported, though most still require validation in paediatric populations (11, 12).

This study aims to evaluate the diagnostic performance of reported novel blood protein biomarkers and biomarker combinations for bacterial infection in a cohort of children aged 16 years and younger, hospitalised with an infection or an inflammatory condition.

## Methods

### Study Design and Setting

This study utilised samples collected as part of a prospective observational matched control study as part of the COVID Warriors study as previously described (13, 14). This prospective observational matched cohort study was conducted at the Royal Belfast Hospital for Sick Children in the UK between May 2020 to January 2023 (ClinicalTrials.gov/NCT04347408). This study was conducted in accordance with the Strengthening the Reporting of Observational Studies in Epidemiology (STROBE) guidelines (15).

### Participants

Children aged 0–16 years admitted to the Royal Belfast Hospital for Sick Children with a suspected infection or inflammatory condition were eligible for inclusion. Participants were recruited via convenience sampling and screened for eligibility at the time of admission by clinical staff. There were no exclusion criteria beyond withdrawal or decline of consent. Participants with insufficient sample availability were excluded from biomarker analysis.

Demographic and clinical data, including presenting features and CRP results, were collected using an electronic case report form. All participants underwent standard clinical care, and additional plasma samples were collected contemporaneous with a routine clinical blood draw. Additional EDTA plasma samples were collected alongside routine phlebotomy required for patient care. Samples were transported to Queen’s University Belfast and frozen at −80°C within six hours of collection.

### Variables and Case Definitions

The outcome of bacterial infection or viral infection was assigned by the research team based on a published phenotyping algorithm (16). The bacterial infection group comprised definite and probable bacterial infections. Inflammatory cases were defined according to established diagnostic criteria, including rheumatic heart disease (Jones criteria), multisystem inflammatory syndrome in children (MIS-C), and Kawasaki disease (Centers for Disease Control and Prevention criteria). The non-bacterial infection group comprised those with viral infection or inflammatory diseases.

### Quantitative variables source and measurement

Biomarkers for measurement in plasma samples were selected from a recent systematic review of blood-based protein biomarkers reported to differentiate bacterial infection with high diagnostic accuracy (12). Biomarker data for Tumour necrosis factor-related apoptosis inducing ligand (TRAIL), Interferon-gamma-inducible protein (IP-10), interleukin-18 (IL-18), Neural cell adhesion molecule 1 (NCAM1), IFN-γ-interferon-gamma (IFN-γ) and E-Selectin were measured in plasma with a bead-based immunoassay, Luminex® Discovery Assay (Bio-techne, Minneapolis, USA). Protein expression levels were measured on a Luminex® 100/200™ analyser, according to the manufacturer’s protocol. Plasma levels of lipocalin-2 (LCN2) and galectin-3-binding protein (LG3BP) were measured via enzyme-linked immunosorbent assay (ELISA) using commercial kits (Quantikine® ELISA kit, Bio-techne, Abingdon, UK). Values that were out of range were replaced by the closest calibrator curve point. Samples were randomised across plates and processed by personnel blinded to case/control status.

The Jackson et al. weighted disease risk scores (DRS) (E-selectin+IL18+NCAM1+LCN2+IFN-γ) and simple DRS (E-selectin+IL18+NCAM1+LCN2+IFN-γ+LG3BP) were calculated as previously described (17). Protein expression values were first log2-transformed. For the weighted DRS, NCAM1, IFN-y, E-Selectin, LCN2 and IL-18 values were multiplied by the published weightings, and the sum of all values was used as the DRS value for each patient. The simple DRS values were calculated by subtracting the sum of the LG3BP, IL-18 and NCAM1 from the sum of LCN2, E-selectin and IFN-y.

The Oved et al. signature (TRAIL+IP-10+CRP) was assessed as per Oved et al., using a multinomial logistic regression model (18). Multinomial logistic regression was performed in R (version 3.6.1) with the nnet package. Predicted probability values for bacterial infection were used as a disease risk score for each participant. Disease risk scores for the novel signature containing LCN2, E-selectin and IFN-y were also generated in this way.

### Statistical analysis

Data were analysed using GraphPad Prism® version 10.0.0 (GraphPad Software, Massachusetts, USA). Normal distribution was determined in all data based on the D’Agostino-Pearson Omnibus normality test. Biomarker measurements were compared between groups using Kruskal-Walis test with Dunn’s multiple comparisons between bacterial infection and other groups. A *P*-value of 0.05 was considered the threshold for significance. Receiver operator curves (ROC) for each biomarker were plotted, and area under the ROC curve (AUC) values were calculated with 95% confidence intervals. AUCs were compared for significant differences using the Delong method in R (version 3.6.1) with the pROC package.

## Results

### Participants

A total of 81 children hospitalised at the Royal Belfast Hospital for Sick Children were screened, 69 of whom gave consent to be included in the case-control study, and 52 children had sufficient sample volume for biomarker work. Participants were classified into three groups based on phenotype: bacterial infection (n=13), viral infection (n=20), and inflammatory condition (n=19). Patient demographics and clinical characteristics are summarised in Table 1.

**Table 1.** Patient characteristics stratified by bacterial, viral and inflammatory groups. IQR; Interquartile range, CRP; C reactive protein, MIS-C; Multisystem Inflammatory Syndrome in Children.

|  | **Bacterial** | **Viral** | **Inflammatory** |
| --- | --- | --- | --- |
|  | n=13 | n= 20 | n= 19 |
| **Age, median years (IQR)** | 7.6 (2.5-10.1) | 1.2 (0.4-5.5) | 8.8 (6.4-13.3) |
| **Males, n(%)** | 7 (53.8) | 11 (55.0) | 9 (47.4) |
| **CRP, median mg/ml (IQR)** | n=13<br>165.9 (69.1-283.0) | n=19<br>10.8 (2.3-24.7) | n=19<br>198.0 (159.0-274.0) |
| **Infection/**<br>**Inflammatory**<br>**condition, n(%)** | Respiratory, 7 (53.8)<br>Appendicitis, 4 (30.8)<br>Bacteraemia, 1 (7.7)<br>Other, 1 (7.7) | Respiratory, 19 (85.0)<br>Encephalitis, 1 (5.0) | MIS-C, 16 (84.2), Kawasaki<br>disease, 1 (10.5)<br>Rheumatic heart disease,<br>2 (10.5) |

### Biomarker results

LCN2, the Oved et al. signature (TRAIL+IP-10+CRP) and two signatures from Jackson et al., a simple signature (E-selectin+IL18+ NCAM1+LCN2+IFN-γ+LG3BP) and a weighted signature (E-selectin+IL18+ NCAM1+LCN2+IFN-γ) were assessed (20, 24-29). This totalled eight novel individual proteins selected.

AUC (95% CI) values for the individual novel biomarkers in distinguishing bacterial infection from viral infection, inflammatory illness, or non-bacterial infection (viral and inflammatory combined) are presented in Figure 1A and Table 2, with box plots shown in Figure 1B. When comparing bacterial and viral groups, only E-selectin and LCN2 were significantly elevated in the bacterial group. Across all eight biomarkers, AUCs for discriminating bacterial from viral infection ranged from 0.542 to 0.926, with E-selectin demonstrating the best performance (AUC 0.926, 95% CI: 0.832–1.020). In comparisons between bacterial and inflammatory groups, NCAM1 and LG3BP were significantly higher in the bacterial group, whereas IP-10 and IFN-γ were significantly elevated in the inflammatory group. AUCs for distinguishing bacterial from inflammatory illness ranged from 0.567 to 0.868, with NCAM1 showing the strongest performance (AUC 0.868, 95% CI: 0.774–0.962). For distinguishing bacterial from non-bacterial infection, AUCs ranged from 0.502 to 0.803, with E-selectin again demonstrating the best discriminatory ability (AUC 0.803, 95% CI: 0.678–0.928).

**Table 2.** AUC values for each biomarker to distinguish bacterial infection from the indicated comparison group.

| **Biomarker** | **Comparison group** | **AUC (95% CI)** |
| --- | --- | --- |
| **NCAM1** | Viral | 0.753 (0.571-0.936) |
|  | Inflammatory | 0.777 (0.615-0.939) |
|  | Non-bacterial | 0.504 (0.339-0.670) |
| **IP-10** | Viral | 0.567 (0.361-0.773) |
|  | Inflammatory | 0.793 (0.630-0.956) |
|  | Non-bacterial | 0.677 (0.515-0.840) |
| **IFN-γ** | Viral | 0.621 (0.430-0.811) |
|  | Inflammatory | 0.742 (0.573-0.912) |
|  | Non-bacterial | 0.680 (0.533-0.827) |
| **E-selectin** | Viral | 0.926 (0.832-1.020) |
|  | Inflammatory | 0.674 (0.486-0.861) |
|  | Non-bacterial | 0.803 (0.678-0.928) |
| **TRAIL** | Viral | 0.542 (0.342-0.742) |
|  | Inflammatory | 0.678 (0.491-0.865) |
|  | Non-bacterial | 0.608 (0.443-0.773) |
| **IL-18** | Viral | 0.603 (0.397-0.810) |
|  | Inflammatory | 0.613 (0.415-0.811) |
|  | Non-bacterial | 0.502 (0.320-0.683) |
| **LG3BP** | Viral | 0.598 (0.399-0.796) |
|  | Inflammatory | 0.744 (0.566-0.923) |
|  | Non-bacterial | 0.569 (0.398-0.739) |
| **LCN2** | Viral | 0.819 (0.661-0.976) |
|  | Inflammatory | 0.607 (0.398-0.816) |
|  | Non-bacterial | 0.716 (0.546-0.885) |
| **CRP** | Viral | 0.927 (0.822-1.00) |
|  | Inflammatory | 0.595 (0.379-0.810) |
|  | Non-bacterial | 0.666 (0.507-0.824) |

**Figure 1.**
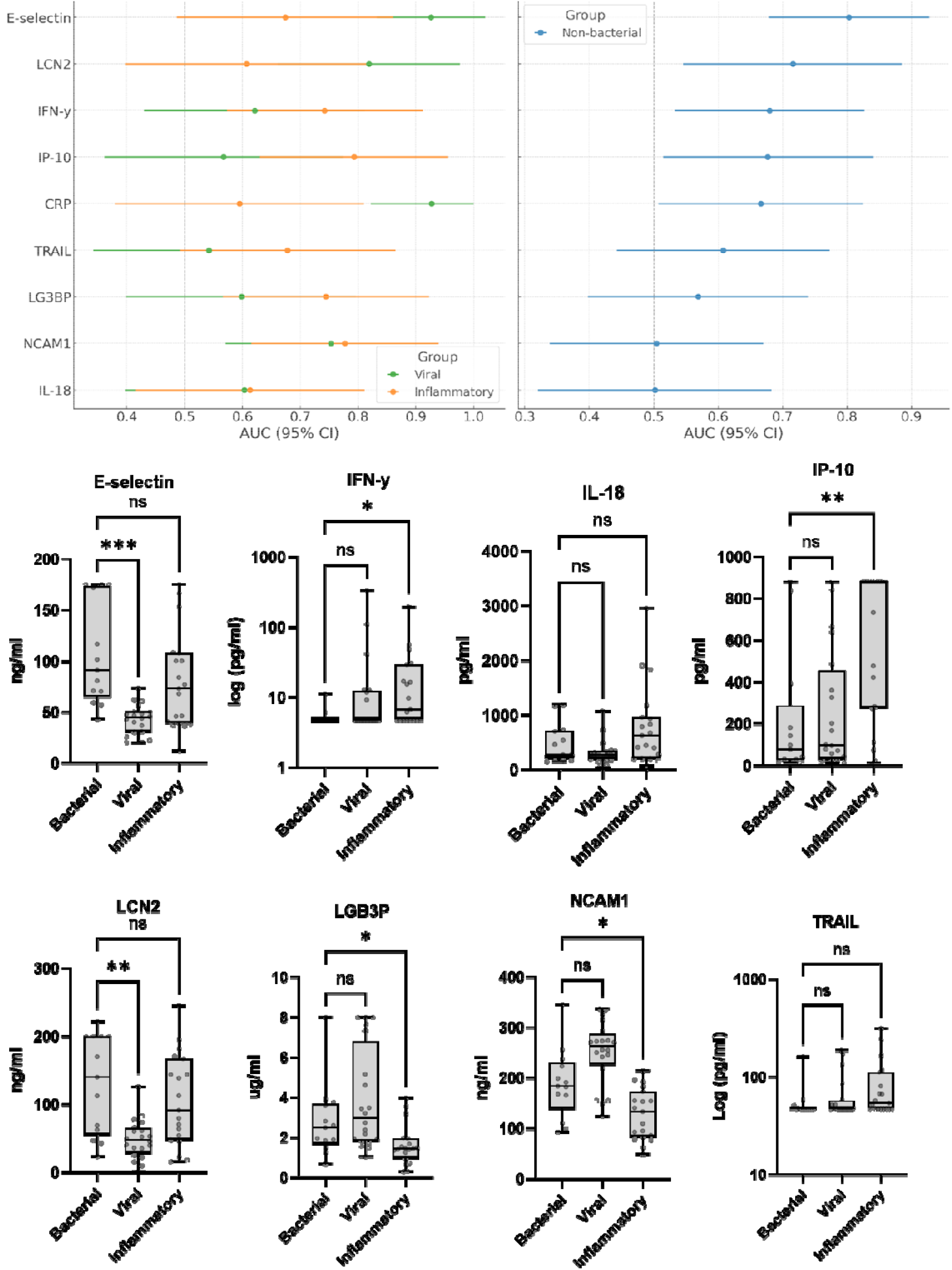
Diagnostic performance of individual biomarkers for distinguishing infection etiologies. (A) Forest plots display the area under the receiver operating characteristic curve (AUC) with 95% confidence intervals (CI) for each biomarker. (B) Boxplot points represent individual data points, horizontal line displays median, boxes are defined by interquartile range, and whiskers extend to the minimum and aximum values. P value annotations show ns>0.05, ^*^<0.05, ^**^<0.01, ^***^<0.001.

Figure 2 shows box plots and ROCs with AUC values for the four biomarker/biomarker signatures selected from the literature: LCN2, the Oved et al. signature, the simple Jackson et al. signature and the weighted Jackson et al signature. All four were significantly different between bacterial versus viral infection, AUCs ranged from 0.819-0.935, with LCN2 having the lowest AUC and the Oved et al signature the highest AUC, 0.935 (95% CI:0.848-1.00). In comparison to CRP alone for bacterial versus viral classification, which had an AUC of 0.927 (0.822-1.00), only the Oved et al. signature had an improved ability to differentiate between bacterial versus viral infection.

**Figure 2.**
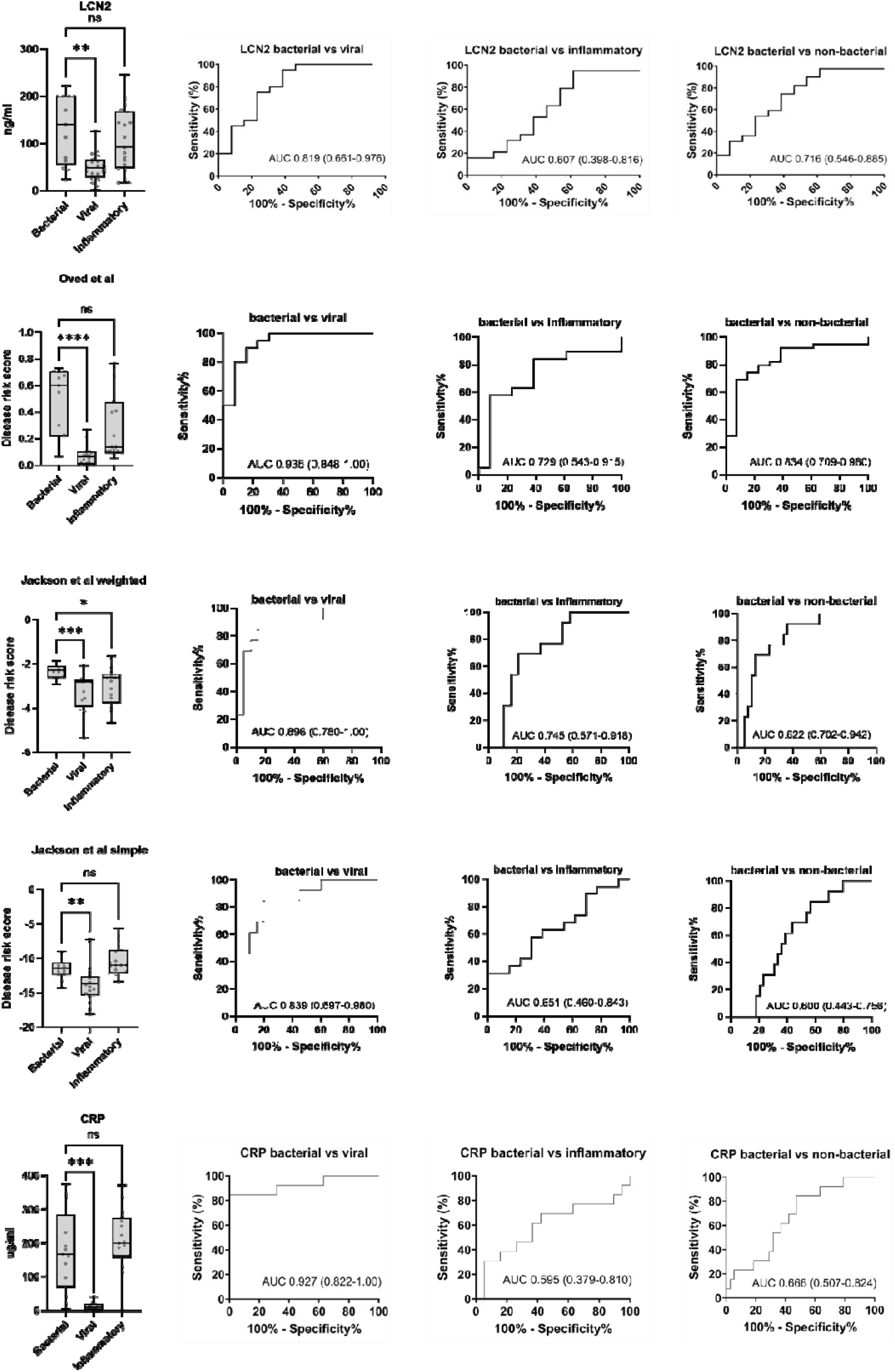
Box plots and ROC curves for biomarkers and signatures identified from the literature. Boxplot points represent individual data points, horizontal line displays median, boxes are defined by interquartile range, and whiskers extend to the minimum and maximum values. P value annotations show ns>0.05, ^*^<0.05, ^**^<0.01, ^***^<0.001. AUC values are displayed on ROCs with 95% confidence intervals in brackets.

For comparison of bacterial versus inflammatory groups, only the weighted Jackson et al. signature was significantly different between groups. AUCs ranged from 0.607-0.745, with the weighted Jackson et al. signature having the highest value, 0.745 (95% CI: 0.571-0.918). In comparison to CRP alone, all signatures had an improved AUC, 0.595 (95% CI: 0.379-0.810) for bacterial versus inflammatory groups. AUCs for the overall comparison of bacterial and non-bacterial infection ranged from 0.600-0.834, with the Oved et al. signature having the highest AUC. In comparison to CRP alone, AUC 0.666 (0.507-0.824), the Oved et al. had significantly improved AUC 0.834 (0.709-0.960), p=0.047.

Given the limited discriminating value of the novel biomarkers and signatures over CRP, we sought to identify a novel combination of host proteins to distinguish bacterial infection from non-bacterial infection. The three proteins with the highest individual AUCs for bacterial vs non-bacterial infection, IFN-y, E-selectin and LCN2 were combined into a signature using multinomial logistic regression. The novel bacterial signature had an improved AUC for bacterial versus viral 0.965 (95% CI: 0.913-1.00) and inflammatory 0.830 (95% CI: 0.685-0.975) compared to CRP (Figure 3). For the comparison of bacterial vs non-bacterial the novel signature had the highest AUC 0.899 (95% CI: 0.817-0.982) of any other single biomarker or signature, and was significantly higher than CRP, p=0.003.

**Figure 3.**
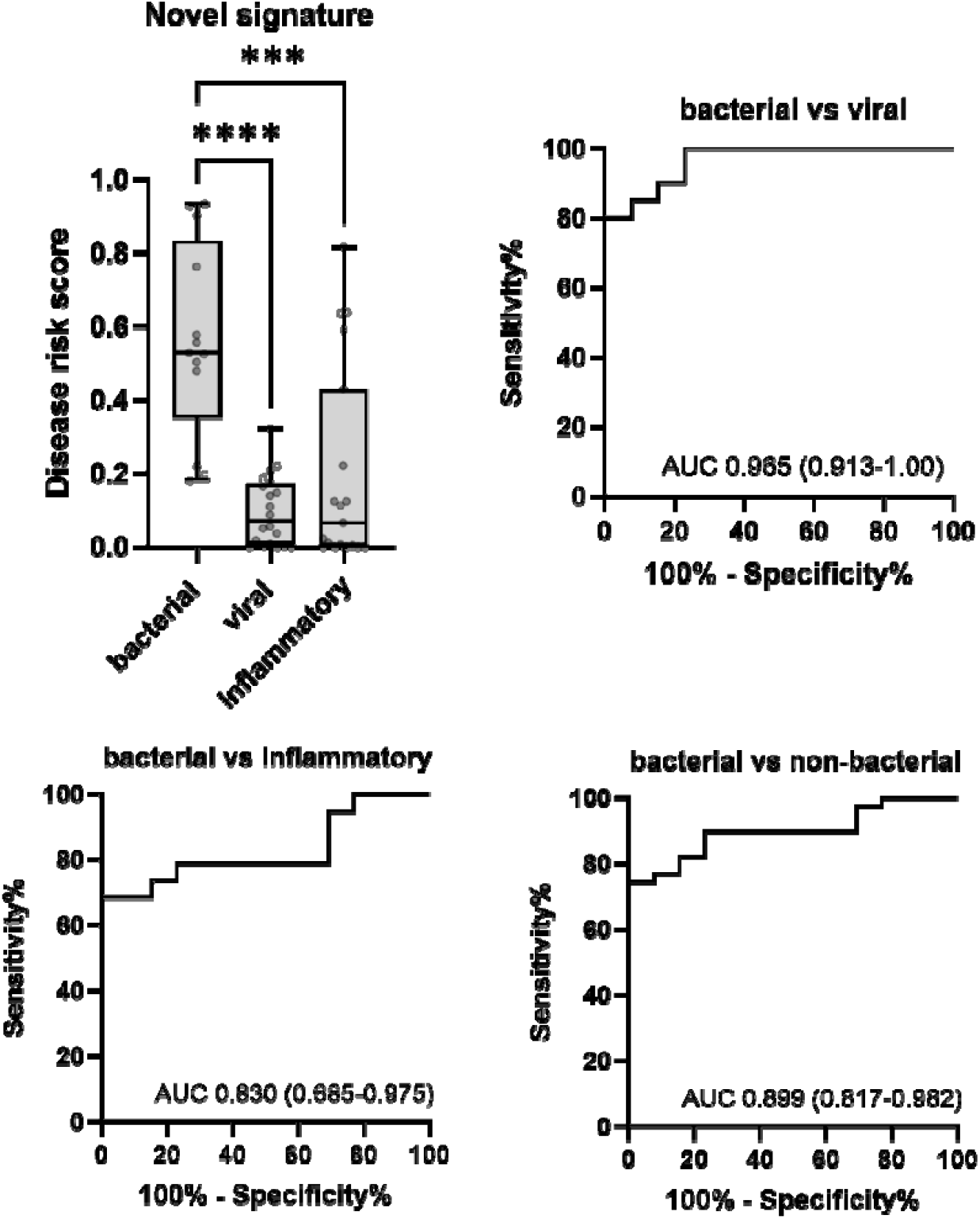
Box plot and ROC curves for a novel biomarkers combination, LCN2, E-selectin and IFN-y. Boxplot points represent individual data points, horizontal line displays median, boxes are defined by interquartile range, and whiskers extend to the minimum and maximum values. P value annotations show ns>0.05, ^*^<0.05, ^**^<0.01, ^***^<0.001. AUC values are displayed on ROCs with 95% confidence intervals in brackets.

## Discussion

This study provides an external validation of novel biomarkers and biomarker signatures, previously reported to have high diagnostic accuracy, for detecting bacterial infection in hospitalised children. Three previously published signatures, Oved et al. (TRAIL+IP-10+CRP), Jackson et al. weighted (E-selectin+IL18+NCAM1+LCN2+IFN-γ), Jackson et al. simple (E-selectin+IL18+NCAM1+LCN2+IFN-γ+LG3BP) and one single biomarker (LCN2) from the literature with promising diagnostic accuracy performance were tested in our cohort (17-20). In total, eight novel biomarkers were evaluated, both individually and as part of multi-protein signatures, in 52 children (≤16 years) hospitalised with either infection or an inflammatory condition.

All three selected signatures, as well as LCN2, distinguished bacterial from viral infections with AUCs ranging from 0.819 to 0.935. Among these, the Oved et al. signature demonstrated the highest diagnostic performance (AUC 0.935) and was the only signature to show higher diagnostic accuracy than CRP (AUC 0.927). This finding is consistent with multiple studies in paediatric cohorts reporting the high diagnostic accuracy of this signature, which is also available on a commercial point-of-care platform from MeMed studies (18, 21-23). The combination of host-response markers, TRAIL (downregulated in bacterial infections), IP-10 (upregulated in viral infections), and CRP (an acute-phase inflammatory marker), likely enhances diagnostic specificity compared with CRP alone. Nevertheless, it is worth noting that the improvement over CRP was modest and, as expected, CRP performed strongly in this cohort of unwell hospitalised children to differentiate bacterial from viral infection.

In contrast, all three biomarker signatures and LCN2 showed poorer discrimination between bacterial and inflammatory conditions than between bacterial and viral infections, with AUCs ranging from 0.607-0.745. This finding is unsurprising, as the original derivation studies did not include patients with inflammatory illnesses. Among the signatures, only the weighted Jackson et al. signature showed a significantly different disease score between bacterial and inflammatory groups (AUC 0.745; 95% CI: 0.571–0.918). Notably, the weighted Jackson signature consistently outperformed the simple version across both bacterial–viral and bacterial–inflammatory comparisons. Although the original authors provided the simplified model due to concerns about potential overfitting in the weighted version, our results suggest that applying weights improved discriminatory performance in this independent cohort (17). Furthermore, the weighted Jackson signature significantly outperformed CRP (AUC 0.595) for bacterial versus inflammatory discrimination, which showed poor performance due to elevated levels in both groups. This aligns with the known limitations of CRP in differentiating bacterial infection from inflammatory syndromes such as MIS-C and Kawasaki disease (24, 25).

We also identified a novel three-protein signature (E-selectin+LCN2+IFN-γ) that significantly outperformed CRP for discriminating bacterial vs non-bacterial infections (inflammatory and viral combined) (P = 0.006). This signature combines markers from distinct immune pathways, endothelial activation, neutrophil activation, and cytokine response, offering improved specificity (26, 27). The inflammatory comparator group predominantly comprised children with MIS-C, reflecting the timing of recruitment during the emergence of this syndrome. As such, generalisability to other inflammatory conditions, including Kawasaki disease, requires further evaluation. Nonetheless, a strength of this study was the inclusion of children with non-infectious causes of fever that often mimic bacterial infection and are frequently diagnosed only after prolonged antibiotic treatment and hospitalisation (28).

This study assessed multiple novel biomarkers of bacterial infection in a vulnerable paediatric population using small sample volumes. Importantly, the cohort included children with non-infectious inflammatory conditions, enabling evaluation of biomarkers across clinically relevant differential diagnoses; however, several limitations should be acknowledged. First, the case–control design may overestimate diagnostic accuracy; however, it represented a pragmatic approach in this context. Second, the relatively small sample size precluded independent training and validation cohorts, and the diagnostic accuracy of the novel signature should be verified in larger studies, ideally including children with co-infections. Third, CRP contributed to the infection classification algorithm, which may bias comparisons in its favour; however, this algorithm is well established, and its use avoided unnecessary sample exclusion (29). Because the Oved et al. signature includes CRP, its diagnostic accuracy may have been inflated.

## Conclusion

This study demonstrates that several novel host protein biomarkers and multi-marker signatures outperform CRP in distinguishing bacterial from viral and inflammatory infections in hospitalised children. The Oved et al. signature was validated as the most accurate tool for bacterial–viral discrimination and is available on a commercial platform, supporting its potential for clinical implementation. For bacterial versus non-bacterial comparisons, particularly in the context of inflammatory illness, the weighted Jackson et al. signature showed the best performance among published tools, highlighting its potential utility in broader clinical cohorts. In addition, our novel three-marker signature (E-selectin+LCN2+IFN-γ significantly outperformed CRP for identifying bacterial infections and warrants external validation in larger, more diverse populations. Together, these findings have important implications for antimicrobial stewardship. Biomarkers that have a higher diagnostic accuracy than CRP could enable earlier and more confident antibiotic de-escalation, reducing unnecessary exposure, drug-related adverse events, hospital-acquired complications, and the development of antimicrobial resistance.

## Study ethics

Ethical approval was provided by the Belfast Health & Social Care Trust Research Governance (Reference 19147TW-SW). Informed consent was obtained for all participants.

## Acknowledgements

The authors thank staff, parents and patients at the Royal Belfast Hospital for Sick Children for the facilitation of sample collection. We particularly thank the Infectious Disease and Paediatric Intensive Care Units staff members, as well as Dr Hannah Bruce Norman and Dr Peter Cosgrove for assistance with sample and data collection.

## Data availability

Data available upon reasonable request.

## AI Use Statement

Artificial intelligence (ChatGPT by OpenAI) was used to assist with improving the clarity, grammar, and readability of the manuscript. All AI-generated content was subsequently reviewed and edited by all authors to ensure scientific accuracy and integrity. No AI tools were used for data analysis or interpretation.

